# Retrospective assessment of SARS-COV2 circulation in two hospital nurseries hosting healthcare workers’ children during lockdown in one of the most affected French area*s*

**DOI:** 10.1101/2020.10.28.20191981

**Authors:** Pauline Penot, Anne Delaval, Fabienne L’Hour, Angélique Grenier, Raya Harich

**Affiliations:** CeGIDD et consultations de maladies infectieuses, centre hospitalier intercommunal André Grégoire, GHT Grand Paris Nord-Est, 56 boulevard de la Boissière, Montreuil, France; Centre Population et Développement (Ceped), Institut de recherche pour le développement (IRD) et Université de Paris, Inserm ERL 1244, 45 rue des Saints-Pères, 75006 Paris, France; Service de Biologie Médicale, Centre Hospitalier Intercommunal Robert Ballanger, GHT Grand Paris Nord-Est, Boulevard Ballanger, 93602 Aulnay sous-bois Cedex France; Direction Qualité Gestion des risques, Centre hospitalier intercommunal André Grégoire, GHT Grand Paris Nord-Est, 56 boulevard de la Boissière, Montreuil, France; Laboratoire de biologie médicale, Centre hospitalier intercommunal André Grégoire, GHT Grand Paris Nord-Est, 56 boulevard de la Boissière, Montreuil, France

**Keywords:** SARS-CoV-2, COVID-19, cluster, nursery, preschool, transmission, children

## Abstract

**Background:** Evidence as to whether childcare and school closure limits the spread of SARS-CoV-2 virus is limited, especially because the role of children in SARS-CoV2 transmission remains unclear.

**Methods:** Between May 29 and July 2, 2020, a retrospective cohort study was conducted among two populations: requisitioned health-care workers and requisitioned staff from hospitals childcare centers, to investigate the virus circulation during lockdown, in a French area of high transmission.

**Results:** The infection attack rate was 6/52 (11.6%) and 8/46 (17.4%) among health-care workers and childcare staff, respectively. An early epidemic occurred among Montreuil s hospital childcare staff, but the parents were not affected (p=0.029). Among Aulnay-sous-bois childcare center, three staff members were infected but none of them was in charge of a child whose parents were infected. Also among the parents of the children they cared for, none developed antibodies. Out of 14 infections, 12 were reliable to a source of transmission, mostly among colleagues.

**Discussion-conclusion:** The assessment of viral circulation among healthcare workers and childcare staff suggests that the children did not contribute to SARS-CoV-2 spread in our setting.

## Background

From March 17 to May 11, 2020, French preschools (6 week-3 year aged children) only remained opened for essential workers’ children. This lockdown had major negative effects on children’s social and mental health, with documented increase of domestic violence and exacerbation of previous social disparities (1) (2). Most European countries have enforced school and preschool closure to mitigate transmission and may do this choice again, as the number of infections is currently increasing all over Europe. It is urgent to assess whether children lockdown might contribute to COVID-19 control. Therefore, we have assessed how SARS-CoV2 has circulated among parents and caregivers in two hospital’s nurseries in an area of important SARS-COV2 circulation when the rest of the population was locked down.

### The TRANSEPS study

We conducted a retrospective closed cohort study among hospital health-care workers who were affected to COVID units and entrusted their children to the hospital’s nurseries during lockdown on one hand, and requisitioned staff from those nurseries on the other hand. The 2 hospitals are located in Montreuil and Aulnay-sous-bois, two cities of Seine Saint-Denis, the French Department with the highest abnormal mortality rate during the first epidemic wave (3). The aim of the study was to assess the viral circulation in those highly exposed adults, connected by 6 week to 3 year aged children, in order to identify possible viral transmission through the infants. Eligible parents had been in charge of COVID patients and had entrusted one or two children to the hospital’s nursery for at least two days between March17 and May 11. Eligible nursery staff had been directly in charge of health workers’ children for at least two days during the same period.

Masks were not required for staff members or children in the two nurseries but barrier measures were established differently: in Aulnay-sous-bois, the staff had access to protective masks and could choose whether to use them or not and the parents could access to the sections provided they wore a mask. In Montreuil, conversely, the childhood professionals had no access to protective masks during lockdown: parents were not allowed to enter and children were entrusted at the entrance to a manager.

Between May 29 and July 2, 2020, all eligible adults were offered participation (see Flowchart on Figure 1). Except for seven, all consented and answered a face-to-face survey requesting details on history of symptoms, contacts, swab for nucleic acid testing, chest tomography and serological testing. Blood specimens were collected and SARS-COV2 IgG detection was done using either Abbott Architect© test (detection rate 1.4 S/C) or Elisa Euroimmun© test (DO sample compared to calibrator: positive result for ratio>=1.1). Any participant with a positive serology at the time of blood sampling was considered as a confirmed SARS-COV2 infection. The infection attack rate (IAR) was defined as the proportion of all participants with confirmed SARS COV-2 infection. This study received ethical approval by the Comité de Protection des Personnes Nord-Ouest II.

**Figure 1:**
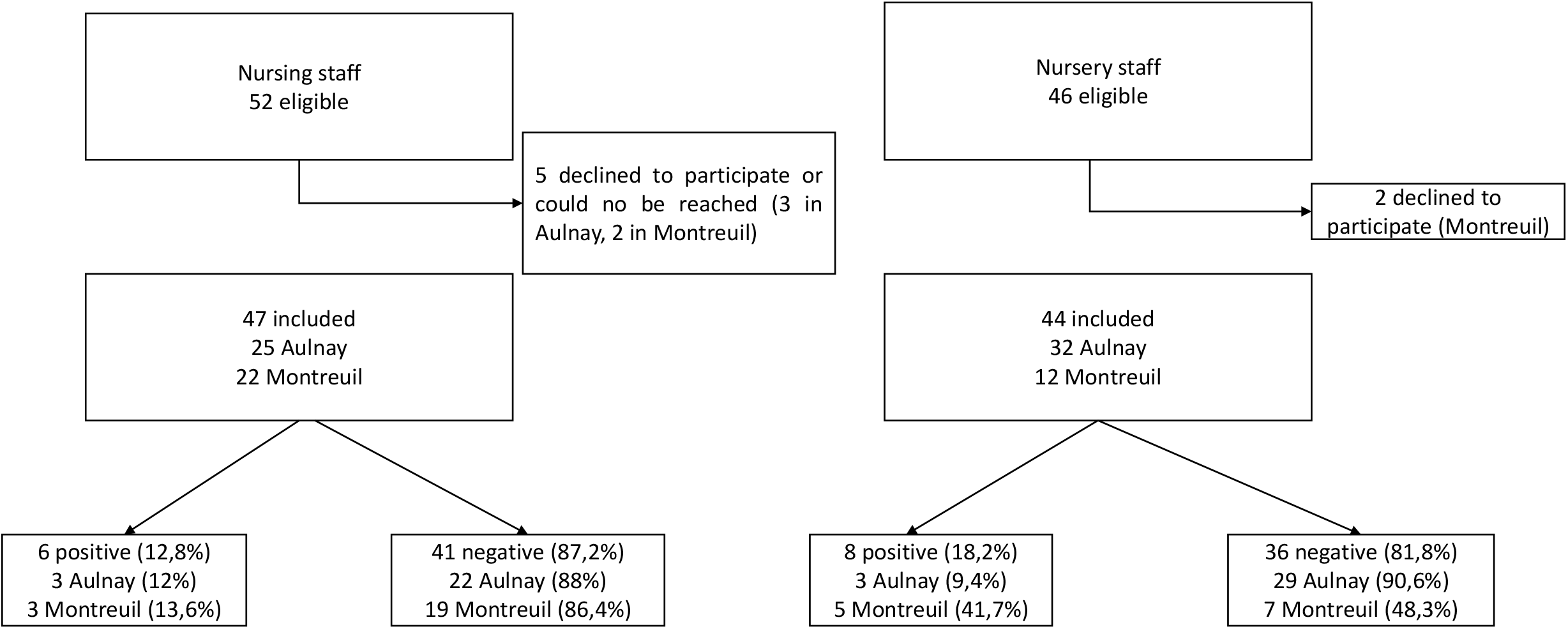
Strategy of recruitment of the study participants, Seine-Saint Denis, France 29 May-2 July 2020.

47 healthcare workers (25 in Aulnay-sous-bois, 22 in Montreuil) and 44 nurseries staff (32 in Aulnay-sous-bois, 12 in Montreuil) were surveyed and sampled (see Table 1). The identification of an outbreak among Montreuil hospital’s nursery staff led to invite incidentally an additional 5 parents whose children were cared for in that nursery during the study period and who were first not eligible as they did not work directly with COVID patients.

**Table 1:**
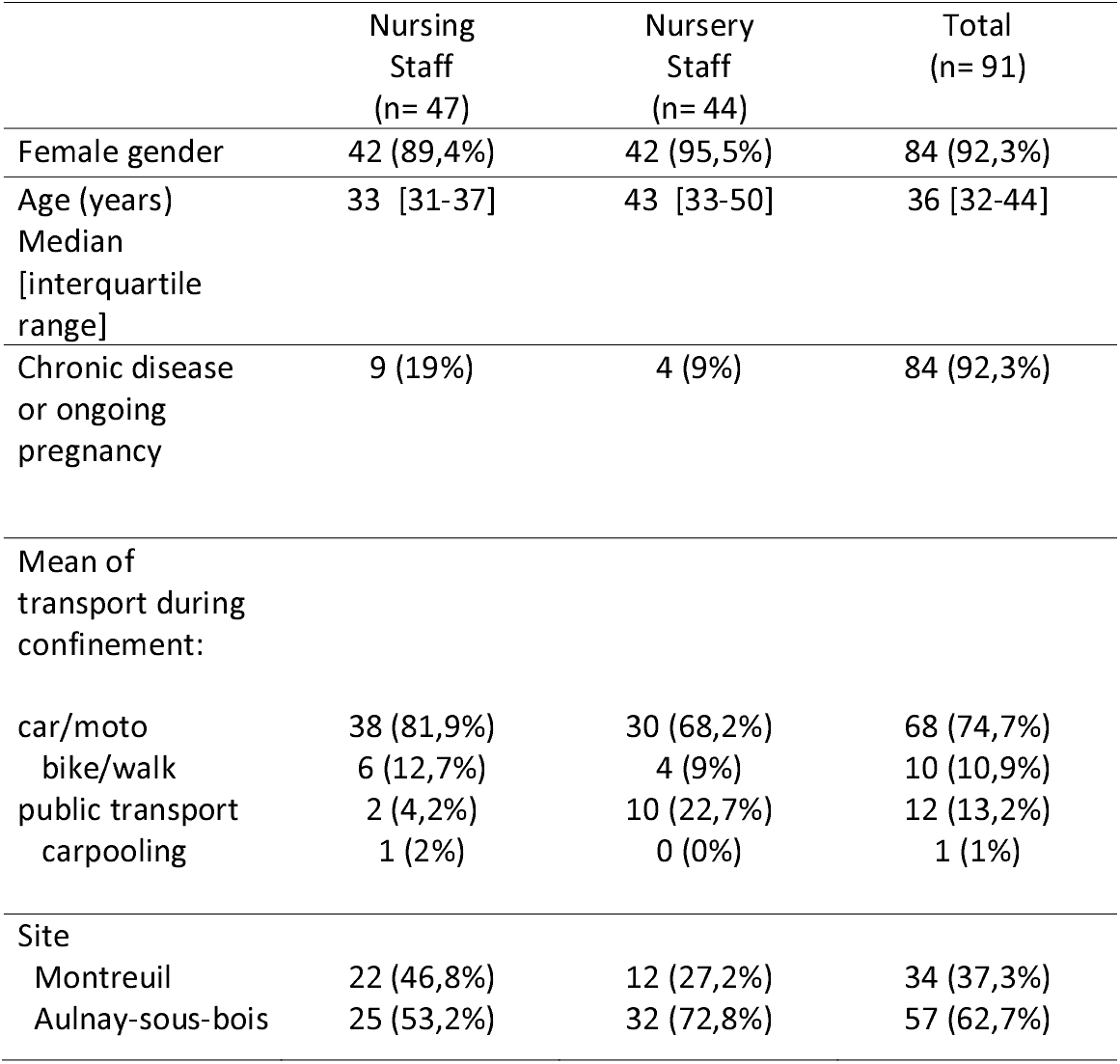
Characteristics of the 91 participants to the TRANSEPS investigation conducted in Seine-Saint-Denis, France, 29 May - 2 July 2020.

|  | Nursing<br>Staff<br>(n= 47) | Nursery<br>Staff<br>(n= 44) | Total<br>(n= 91) |
| --- | --- | --- | --- |
| Female gender | 42 (89,4%) | 42 (95,5%) | 84 (92,3%) |
| Age (years)<br>Median<br>[interquartile<br>range] | 33 [31-37] | 43 [33-50] | 36 [32-44] |
| Chronic disease<br>or ongoing<br>pregnancy | 9 (19%) | 4 (9%) | 84 (92,3%) |
| Mean of<br>transport during<br>confinement: |  |  |  |
| car/moto | 38 (81,9%) | 30 (68,2%) | 68 (74,7%) |
| bike/walk | 6 (12,7%) | 4 (9%) | 10 (10,9%) |
| public transport | 2 (4,2%) | 10 (22,7%) | 12 (13,2%) |
| carpooling | 1 (2%) | 0 (0%) | 1 (1%) |
| Site |  |  |  |
| Montreuil | 22 (46,8%) | 12 (27,2%) | 34 (37,3%) |
| Aulnay-sous-bois | 25 (53,2%) | 32 (72,8%) | 57 (62,7%) |

Surprisingly, the IAR was not higher among the COVID units’ healthcare workers (IAR 11.1% in Montreuil and 12% in Aulnay-sous-bois) than among the nurseries’ professionals (41.7% in Montreuil and 9.4% in Aulnay-sous-bois). An outbreak has even occurred among the nursery’s staff in Montreuil without affecting the parents (p=0.029).

### Description of Montreuil’s nursery outbreak and viral circulation among parents

In Montreuil, four childcare assistants belonging to the usual team (cases 1 to 4) and a staff member who came as backup during lockdown (case 5) had positive SARS-COV2 antibodies.

The case “Montreuil Nursery 1” (MN1) had prolonged flu-like symptoms from February 3. Her partner had COVID-compatible symptoms from March 17.

MN2’s symptoms started on March 10, MN3’s and MN4’s on March 13. On Mars 15, MN4 was hospitalized for respiratory distress with confirmed SARS-COV2 diagnosis.

MN1, 2 and 3 worked in the same unit until lockdown reduced the nursery to a single unit, dedicated to requisitioned healthcare worker’s children. They had daily lunch with MN4.

MN5 developed typical SARS-COV2 symptoms later, on April 6. He was in charge of interfacing with the parents at the admission and departure of the children.

A sixth childcare assistant developed flu-like symptoms by late February, was on sick leave during the study period, and therefore not included, despite SARS-COV2 positive antibodies.

Three parents had positive serology: a nurse from the emergency room (case “Montreuil Parent1” or MP1) developed symptoms on March 21, with microbiological confirmation on March 28. The contaminator was probably her work-team, diagnosed by RT-PCR for symptoms on March 14. MP1’s daughter has been kept out of the nursery from her mother’s symptoms onset (March 21) until April 15.

A physician who worked in the Infectious Disease ward during the outbreak had 2 successive viral episodes: a flu-like syndrome on February 10, during her maternity leave, solved before her daughter’s first stay at the nursery, then an isolated cough from March 21, 2 weeks after close contact with 2 infected colleagues. RT-PCR performed on March 25 was negative. Two chickenpox-like vesicles (blisters?) compatible with dermatological COVID lesions appeared on her infant’s leg on April 15 (4).

A nurse assistant (MP3) developed SARS-COV2 antibodies without any history of symptoms. From April 21, she was posted in the COVID geriatrics department, where she shared resting time and room with four colleagues whose infection was retrospectively confirmed by PCR. One in particular was the nurse she worked in tandem with: they shared a 12 hours day of work while this nurse was symptomatic, before her eviction for positive PCR.

It is noteworthy that a 2-year-old child who attended the nursery from March 17 to May 5 was hospitalized on May 13 for a Kawasaki-like vasculitis. Repeated microbiological as well as serological tests remained negative for him and for both of his parents.

### Viral circulation among nursery staff and parents’ from Aulnay-sous-bois Hospital

In the second nursery, three childhood professionals and three parents from COVID units were infected with SARS-COV2.

Two childhood professionals were in close contact together and both exposed on early March to relatives clinically suspected of COVID, while the third had no other risk factor than daily use of public transport.

The three positive parents had all presented typical COVID symptoms, at different periods: March 26, April 4 and 15. They have all reported prior contact with a RT-PCR confirmed case, in a timeframe compatible with viral transmission: two with colleagues, the third in her private sphere. The childhood professionals in charge of this parent’s children remained negative.

## Discussion

We examined transmission among COVID exposed health-care workers and among childcare professional in charge of their children and found no evidence for onward transmission from one group to the other trough the infants. Our results are consistent with other findings on early childcare settings, where child to staff’s transmission appeared unlikely to have occurred (5) (6) (7) (8) (9). Out of ten Australian preschools monitored for a staff member or a child identified with COVID infection, forward transmission was only documented in one, where the index case was an adult (10) but in Utah, a 8 month old baby may have transmitted preschool-acquired COVID to both parents (11). Overall, SARS-COV2 infections and outbreaks appear uncommon across early year settings and involved mostly staff members (12) (13). One possible explanation for this low rate of onward transmission from children is the high frequency of asymptomatic infections in this population, as high viral loads were reported in pharyngeal swabs from symptomatic children (14) including 0-5 year old ones (15).

The relatively low IAR among the parents goes against our initial hypothesis of higher risk for directly exposed healthcare workers. This IAR is consistent with the global IAR among Montreuil’s (14.8 %) and Aulnay-sous-Bois’ (13%) hospital staff measured on June 25 and pleads for the efficiency of barrier measures, especially as most of the adults cases were linkable to a colleague’s infection.

## Data Availability

All data are available

Despite scare data on age-specific transmissibility, young children appear unlikely to initiate or propagate SARS-COV2 outbreaks: it seems reasonable to allow them to resume activities and social life.

We declare no competing interest and no funding for this work.

## Acknowledgments

We thank the two nurseries’ teams, especially the directors (Helen Giles, Isabelle Legrand, Yaël Sibony, Leslie Negrit) and the Maison Bleue^©^ group (operator for the Montreuil hospital nursery). We also thank Sandrine Dekens, Anne-Laurence Doho, Valérie Millul, Amélie Jean, as well as all the participants (parents and childhood professionals).

## References

1. UNESCO. Impact du COVID sur l’éducation [Internet]. [cited 2020 Aug 13]. Available from: https://fr.unesco.org/covid19/educationresponse

2. Van Lancker W, Parolin Z. COVID-19, school closures, and child poverty: a social crisis in the making. Lancet Public Health. 2020;5(5):e243–4.

3. Gascard N, Kauffmann B, Labosse A. 26 % de décès supplémentaires entre début mars et mi-avril 2020□: les communes denses sont les plus touchées [Internet]. Insee; 2020 May [cited 2020 Sep 15]. Report No.: 191. Available from: https://www.insee.fr/fr/statistiques/4488433

4. Wollina U, Karadağ AS, Rowland-Payne C, Chiriac A, Lotti T. Cutaneous signs in COVID-19 patients: A review. Dermatol Ther. 2020 May 10;e13549.

5. Heavey L, Casey G, Kelly C, Kelly D, McDarby G. No evidence of secondary transmission of COVID-19 from children attending school in Ireland, 2020. Euro Surveill Bull Eur Sur Mal Transm Eur Commun Dis Bull. 2020;25(21).

6. Yung CF, Kam K-Q, Nadua KD, Chong CY, Tan NWH, Li J, et al. Novel coronavirus 2019 transmission risk in educational settings. Clin Infect Dis Off Publ Infect Dis Soc Am. 2020 Jun 25;

7. Danis K, Epaulard O, Bénet T, Gaymard A, Campoy S, Botelho-Nevers E, et al. Cluster of Coronavirus Disease 2019 (COVID-19) in the French Alps, February 2020. Clin Infect Dis Off Publ Infect Dis Soc Am. 2020 28;71(15):825–32.

8. Fontanet A, Grant R, Tondeur L, Madec Y, Grzelak L, Cailleau I, et al. SARS-CoV-2 infection in primary schools in northern France: A retrospective cohort study in an area of high transmission [Internet]. Infectious Diseases (except HIV/AIDS); 2020 Jun [cited 2020 Aug 13]. Available from: http://medrxiv.org/lookup/doi/10.1101/2020.06.25.20140178

9. Dub T, Erra E, Hagberg L, Sarvikivi E, Virta C, Jarvinen A, et al. Transmission of SARS-CoV-2 following exposure in school settings: experience from two Helsinki area exposure incidents. [Internet]. Infectious Diseases (except HIV/AIDS); 2020 Jul [cited 2020 Aug 13]. Available from: http://medrxiv.org/lookup/doi/10.1101/2020.07.20.20156018

10. Macartney K, Quinn HE, Pillsbury AJ, Koirala A, Deng L, Winkler N, et al. Transmission of SARS-CoV-2 in Australian educational settings: a prospective cohort study. Lancet Child Adolesc Health. 2020 Aug;S2352464220302510.

11. Lopez AS, Hill M, Antezano J, Vilven D, Rutner T, Bogdanow L, et al. Transmission Dynamics of COVID-19 Outbreaks Associated with Child Care Facilities — Salt Lake City, Utah, April–July 2020. MMWR Morb Mortal Wkly Rep. 2020 Sep 18;69(37):1319–23.

12. Ismail SA, Saliba V, Lopez Bernal JA, Ramsay ME, Ladhani SN. SARS-CoV-2 infection and transmission in educational settings: cross-sectional analysis of clusters and outbreaks in England [Internet]. Public and Global Health; 2020 Aug [cited 2020 Sep 14]. Available from: http://medrxiv.org/lookup/doi/10.1101/2020.08.21.20178574

13. European Center for Disease Control. COVID-19 in children and the role of school settings in COVID-19 transmission. Stockholm: ECDC; 2020 Aug.

14. Jones TC, Mühlemann B, Veith T, Biele G, Zuchowski M, Hoffmann J, et al. An analysis of SARS-CoV-2 viral load by patient age [Internet]. Infectious Diseases (except HIV/AIDS); 2020 Jun [cited 2020 Aug 20]. Available from: http://medrxiv.org/lookup/doi/10.1101/2020.06.08.20125484

15. Heald-Sargent T, Muller WJ, Zheng X, Rippe J, Patel AB, Kociolek LK. Age-Related Differences in Nasopharyngeal Severe Acute Respiratory Syndrome Coronavirus 2 (SARS-CoV-2) Levels in Patients With Mild to Moderate Coronavirus Disease 2019 (COVID-19). JAMA Pediatr [Internet]. 2020 Jul 30 [cited 2020 Sep 1]; Available from: https://jamanetwork.com/journals/jamapediatrics/fullarticle/2768952

